# Self-help and Prescription Therapies for Menstrual Burden in a University Population

**DOI:** 10.64898/2025.12.30.25343226

**Authors:** SE Zimberg, AM Castejon, N del Mazo, C. Murphy

## Abstract

**Background:** It is known that cyclic menstruation imposes significant burdens on a large portion of the female population with a plethora of symptoms, including pelvic pain, dysmenorrhea, menorrhagia, and psychological distress. The effects are widespread, impacting the quality of life, ability to work and function, finances, and education. For the most part, previous studies have been surveys administered through social media channels such as pelvic pain and endometriosis societies, which, by definition, introduce bias. There is a need to evaluate the female population regarding female health issues in a less biased manner to assess the actual disease burden and response.

**Methods:** A cross-sectional, IRB-approved study of the Nova Southeastern University undergraduate (2022) and graduate female population cohort was conducted using a self-administered, anonymous survey to assess the prevalence of menstrual pain and disability (menstrual burden). This included the use of prescription and non-prescription medications, self-help interventions, and cannabis use (either medicinal or recreational) for symptoms associated with menstruation. All students who identify as female at the university were invited via campus email to participate in this survey, hosted securely on the SurveyMonkey website. One initial invitation was sent, followed by two reminder invitations at 2-week intervals. A descriptive analysis was performed.

**Results:** 14,024 email invitations were sent to the entire university population that identified as female to the registrar, with a response rate of 15.8% (similar to the response rate in most university surveys at this institution). 18.23% of the cohort reported bleeding that restricted the student’s ability to function, and 13.2% were so severely affected by menstrual symptoms that bed rest was required to cope adequately. Primary symptoms of bloating/swelling/constipation were reported in over 77%, mood swings/moodiness in approximately 74%, and pelvic or back pain was noted in over 70% of the respondents. In an assessment of treatment regimens, approximately 80% used over-the-counter medications, 55% reported using heating pads, 25% used oral contraceptive products, 29.6% engaged in exercise or meditation, and fully 14.76% used cannabis in its various forms as treatment adjuncts (in addition to other regimens). In the evaluation of multiple efficacies, the respondents reported that 72% of those that used OTC medications for relief found them very or moderately effective, 66.2% of those that used a heating pad found it very or moderately effective, 54.2% of those that used exercise and meditation found it very or moderately effective, and cannabis was found to be very or moderately effective by 82% of the cohort that reported using it as an adjunct. Unexpectedly, there were some racial/ethnic differences in disease burden, the types of treatment modalities accessed, and perceived effectiveness. There were minimal differences between age groups.

**Conclusion:** Menstruation in female college students represents a significant challenge for 40.2% of this South Florida population, causing moderately severe to extremely severe symptoms. This study supports previous findings by Schoep^2^ on the impact of menstruation on the quality of life of Dutch women and by Munro’s global systematic review^3^, which documented a significant menstrual burden and its implications for education. We found a high burden among college women when the entire female university population was invited to participate in this survey. This study’s emphasis on the menstrual burden and its impact on quality of life expands on previous studies. Our results should pave the way for a policy review of how the menstrual burden is approached in university settings, particularly regarding efforts to encourage gender equality.

## Introduction

As of 2021, there were approximately 2 billion girls and women of reproductive age (ages 14 to 50) worldwide.^1^ Monthly menstruation imposes significant burdens with a plethora of symptoms, including pelvic pain, dysmenorrhea, menorrhagia, and psychological distress affecting a large portion of this population. The effects are widespread, impacting the quality of life, ability to work and function, finances, and education. A recent Dutch study (Schoep et al. 2019)^2^ explored the impact of menstrual symptoms among 42,879 subjects who were reached through social media over a several-month period in 2017, using a cross-sectional survey. The mean age of respondents was 28.7 years (SD 8.6), and 96.8% were Dutch. The most common complaints were abdominal pain during the period (87.4%), perimenstrual psychological complaints (77.3%), and tiredness (70.7%), with heavy bleeding, back pain, and headache accounting for 53.7%, 59.2%, and 56.2% respectively, significantly affecting the respondents’ quality of life. More than 33% of the women were limited in their functioning during menstruation, and the annual costs surpassed $2,000 (US) per patient, primarily secondary to lost work and productivity.

To contextualize the effects of menstruation on university students, Munro et al. (2021)^3^ conducted a systematic review (83 studies included) of the respondents’ experiences and the impact on education worldwide. In general, they reported negative experiences primarily from dysmenorrhea, difficulties containing the menstrual flow, and the shame and stigma associated with the flow. Most respondents reported missing school, poor social engagement, and decreased academic performance monthly. Some societies (very few) reported positive experiences in reinforcing womanhood, though many cited a lack of sanitary utilities to accommodate personal care of the menstrual flow. Because studies have indicated that improved education is linked to stronger economies in which women work and to improved social and family standing, documenting the barriers contributing to their failure is a public policy imperative.^4^

One of the common disease states impacting menstrual burden is endometriosis. Endometriosis is a disease affecting up to 10% of reproductive-age women, sometimes causing diffuse and often severe pelvic pain and dyspareunia. The prevalence can reach 50% in women with a history of infertility.^5^ The disease is defined as the presence of endometrial tissue located outside the uterine cavity. It can involve the ovaries, bladder, rectum, pelvic peritoneum, and even distant areas such as the bowels and lungs. The varying locations are responsible for the multitude of disparate symptoms (primarily pain) and infertility.^6^ The global endometriosis market is forecast to reach $2.42 billion by 2026, encompassing medical, holistic, and surgical treatments. The market is projected to grow significantly during the forecast period, with one notable factor being the rising incidence of the condition.^7^ Treatment for the disease is dependent on the severity of the disease, symptoms, and the patient’s desire to have children. Medication is usually tried first when pain is the primary problem; however, medical treatment (such as Non-Steroidal Anti-Inflammatory Drugs - NSAIDs) is often insufficient to treat the symptoms, and therefore, hormonal and/or surgical intervention is required.^8^

In the general population, it is estimated that between 16% and 91% of women experience pelvic pain and dysmenorrhea (painful menses) regularly.^9^ There have been several exploratory studies in the recent past, notably in Europe, South Africa, Australia, and New Zealand, regarding self-treatment and management of endometriosis and menstrual-associated pelvic pain. Armour et al. (2019) reported on self-management strategies among Australian women with endometriosis.^5^ Of the 484 usable responses documenting self-management of symptoms, 70% used heat (heating pads), 68% used bodily rest, 47% used meditation and breathing, and 14% and 13% used alcohol and cannabis, respectively. Of all the various interventions, women in this study rated cannabis with tetrahydrocannabinol (THC) as the most effective adjunctive therapy, with 56% able to reduce their endometriosis medications by more than 50%. Only 1/3 of CBD (cannabidiol) users achieved a 50% reduction of medications, and the effectiveness of the other interventions ranked far behind cannabis and CBD.^5^ Recent systematic reviews of the effectiveness of dietary interventions on endometriosis and pelvic pain showed positive effects. Still, the assessment of bias risk was high.^10^ Similarly, though women widely practice changes in lifestyle for the treatment of menstrual pain and endometriosis, the world’s scientific literature is conflicting on its value.^11^ Several studies based on surveys indicate that cannabis with THC, more so than CBD, may also be helpful in the treatment of pain associated with endometriosis.^12,13^ Additionally, recent reviews have focused on the endocannabinoid system as a primary nexus of the pain and other symptoms associated with menstrual pain and endometriosis: nociceptive, inflammatory, and neuropathic. This implies that cannabis may play a significant role in the treatment of endometriosis with fewer side effects compared to conventional treatment and possibly delay the need for invasive management (Ahmad 2021, p. 344-345), and is complemented by the expression of cannabinoid receptors, hormones, enzymes, and ligands.^13,14^ The endocannabinoid system (ECS) modulation then presents a reasonable strategy to combine all the factors above to treat the symptoms of menstrual pain and endometriosis. Cannabis is a readily available modulator that can then be used as an adjunct treatment.

The purpose of this study is to evaluate the current state of menstrual burden and adjunctive therapeutic modalities in a college-age cohort, including cannabis, to ameliorate the symptoms of menstruation, menstrual pain, and endometriosis, and to assess the perceived effectiveness of said interventions. Most previous studies have used social media-based tools through websites dedicated to menstrual pain and endometriosis, which, by design, can introduce an inherent bias into the findings. A 2021 study from six Fellowship of Minimally Invasive Gynecologic Surgery sites (AAGL) documented that patients with gynecologic pain were more likely than those without pain to use social media and the internet to understand and manage their condition. Patients with pain engaged in and trusted social media at a higher level, with engagement rising directly with the menstrual burden.^15^ Additionally, the placebo effect and media expectations of efficacy weigh heavily in studies assessing the effectiveness of cannabis therapy for pain.^16^ Although bias can never be eliminated, using an entire university cohort that identifies as female should mitigate much of it and is a unique approach among current studies. We documented a high menstrual burden on the female university population when the entire female university college student body was invited to respond, rather than focusing on pain and endometriosis groups from social media platforms.

## Methods

### Study Design

This is a cross-sectional study of the Nova Southeastern University undergraduate and graduate female population cohort using an anonymous survey to collect data about the prevalence of menstrual pain and disability, as well as the use of various treatment modalities, including cannabis (either medicinal or recreational) for pelvic pain associated with cyclic menstruation and endometriosis. The study was approved by the Nova Southeastern University Institutional Review Board (Study on Women’s Menstrual Symptoms and Treatment– NSU IRB Protocol Number 2021-544). It also explored the dosage forms and formulations used, as well as the perceived effectiveness of the different modalities in relieving symptoms. This was a 12-question survey taking less than 5 minutes (average 3 minutes) to complete online. All students who identified as female to the university registrar were invited via campus email to participate in this survey, hosted securely on SurveyMonkey. One initial invitation was sent, followed by two reminders sent at 2-week intervals. The survey stopped collecting responses on 2/15/22.

NSU had 20,888 students enrolled for the 2021-2022 academic year (of whom 6,314 were undergraduates, and 14,574 were graduate students). The number of graduate and undergraduate students varies significantly on the university website. Of the total enrollment (20,888), 70% were female, and 30% were male (2.3 to 1 ratio).^17^ The racial/ethnic breakdown was 6,447 Hispanic, 6,105 White, 3,676 Black or African American, 1,752 Asian, 938 International, 1,057 Unknown, 577 Multi-ethnic, 24 Native American or Pacific Islander. Of note, the NSU student population does not mirror the US general population in terms of race and ethnicity, though practical conclusions can still be drawn from this cohort.^18^ The survey is included as Addendum 1 at the end of this manuscript after the references. The enrollment statistics were from self-identification to the university registrar.

### Inclusion/Exclusion Criteria

All subjects were included if they were female graduate or undergraduate students at NSU between the ages of 18 and 50 who responded to the survey. As an enrolled NSU student, it was assumed that the subjects had access to a computer and basic English proficiency. No one who responded was excluded from the analysis.

### Recruitment and Data Collection

Subjects were recruited via an online letter explaining the purpose of the study, sent via the university email system to graduate and undergraduate students, with an email link to a secure SurveyMonkey data collection form. Response reminders were sent twice to encourage participation. Results were collected anonymously using SurveyMonkey forms that offered secure socket layer encryption. The survey link was closed after two months, and the results were analyzed. A contact email was provided to subjects for questions, and options were offered to receive an executive summary after publication of the results. Results are stored securely on the Survey Monkey site. No personal identification was collected; only general demographics were collected.

### Outcomes

This survey provides insight into the prevalence and degree of menstrual burden, menstrual symptom treatment preferences, and cannabis use as an adjunct for treating menstrual pain in female college students. It also assesses the prevalence of cannabis inhalation (smoking and vaping), edibles (gummies, foods, beverages), topical (cream, gel, patch), and other (spray, suppository) for treatment. Formulation data were collected on THC-dominant, CBD-dominant, and balanced THC/CBD mix popularity, and the perceived efficacy of various adjuncts noted in previous papers, including cannabis, for relieving symptoms, such as pelvic pain, cramps, bloating, and fatigue. Demographic information was collected to assess racial/ethnic differences in use and the perceived effectiveness of the various modalities used for relief.

## Results

An invitation to complete the 12-question survey (with an embedded link) was sent to all graduate and undergraduate students who identify as female at Nova Southeastern University (n=14,024). The university sent 14,024 invitations from its separate enrollment records, which are 1.7% higher than what is published on the university website. This was over a 7-week span in January and February of 2022. Reminder letters were sent two weeks and four weeks after the initial request. There were 2,216 responses for a total response rate of 15.8%. A review of the University’s response to its annual surveys has hovered around 16% (personal communication, NSU Office of Institutional Effectiveness), which is comparable to this survey experience. There was an immediate response on the first day after the email invitation was sent, then a rapid drop-off over the following week, as seen in Figure 1. The initial response was 7.9% on the day following the initial invitation, followed by 4.2% on the day following the first reminder, and 3% on the day following the second reminder, as seen below.

**Figure 1.**
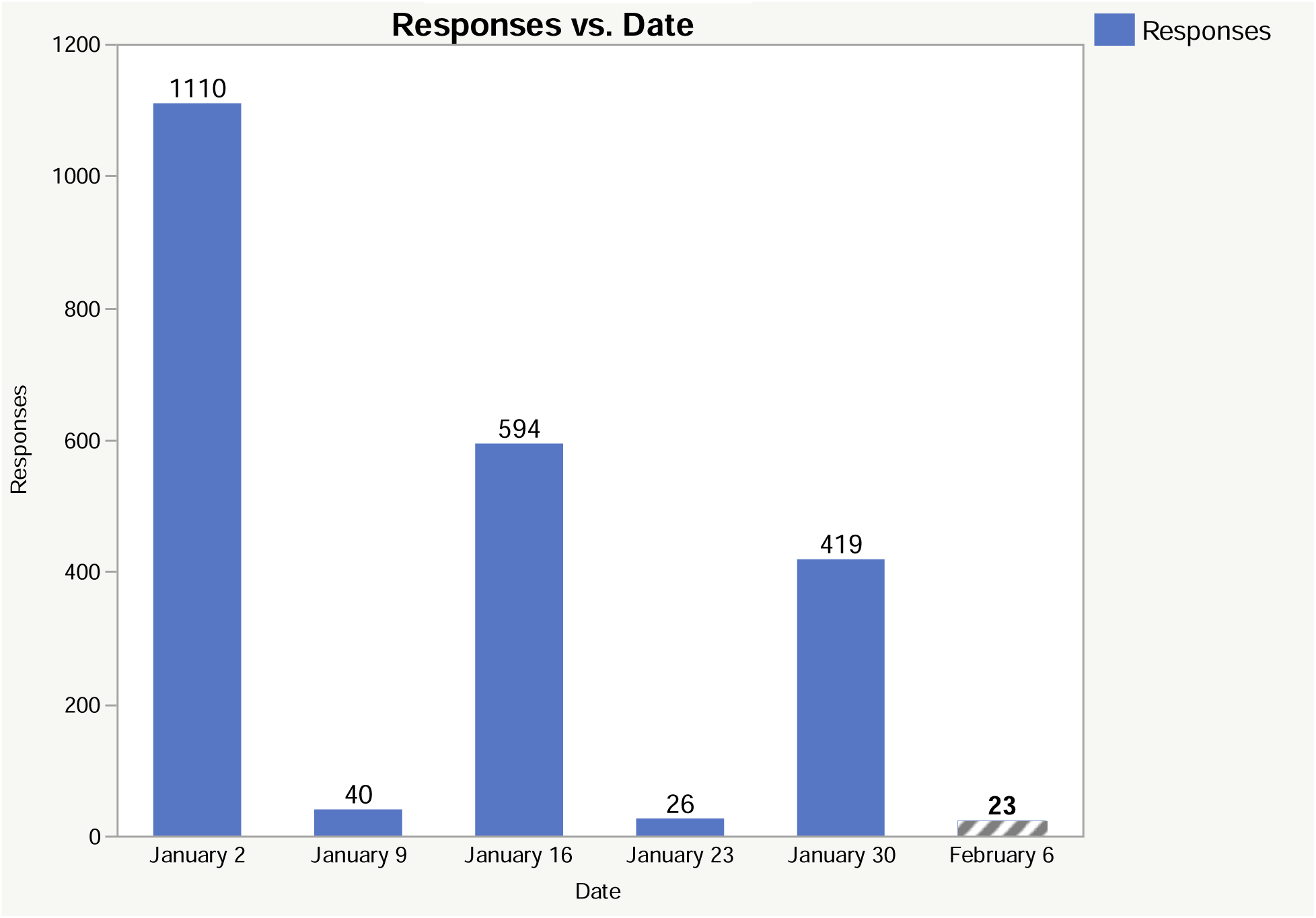
Display of survey responses by date, with January 2 corresponding to the initial email survey request, January 16^th^ to the second reminder, and January 30^th^ to the third reminder. The responses a week out from each are also listed. 4 responses came in late, bringing the total to 2216.

### Demographics

The responses were divided into graduate (59.38%) and undergraduate (40.61%), as shown in Table 1 below.

**Table 1.** Academic Level of Respondents. Breakdown of academic levels among all respondents. Nova Southeastern University currently has 6,314 undergraduate and 15,531 graduate students (the total graduate varies on the university website from 14,574 to 15,531), making up the total student body for a 1 to 2.3 ratio (undergraduate to graduate). Survey responses were 40.61% undergraduate and 59.38% graduate, for a 1:1.46 response ratio. The differences between the total responses in Figure 1 and Table 1 reflect variations in responses to the various questions.

| **Education Level of Respondents** | **Raw Response Number** | **Percent of Responses (n=2238)** |
| --- | --- | --- |
| Undergraduate Freshman | 275 | 12.29% |
| Undergraduate Sophomore | 199 | 8.89% |
| Undergraduate Junior | 221 | 9.87% |
| Undergraduate Senior | 214 | 9.56% |
| 1<sup>st</sup> Year Graduate School | 475 | 21.22% |
| 2<sup>nd</sup> Year Graduate School | 418 | 18.68% |
| 3<sup>rd</sup> Year Graduate School | 210 | 9.38% |
| 4<sup>th</sup> Year Graduate School | 118 | 5.27% |
| Beyond 4<sup>th</sup> Year (PG5-PG7) | 108 | 4.83% |
| **Total** | **2,238** | **99.99%** |

From an age perspective, 81.96% of the respondents were under 30. This reflects the nature of the university undergraduate and graduate students, with younger respondents making up the bulk. The breakdown is shown in Table 2.

**Table 2.** Age Distribution of Respondents. The age distribution of the respondents. 81.96% were age 30 or younger. There are 22 fewer respondents to this query than to the Academic Level question in Table 1.

| **Age Range of Respondents** | **Raw Response Number** | **Percent of Responses (n=2216)** |
| --- | --- | --- |
| 17 to 20 | 579 | 26.13% |
| 21 to 25 | 838 | 37.82% |
| 26 to 30 | 399 | 18.01% |
| 31 to 35 | 146 | 6.59% |
| 36 to 40 | 90 | 4.06% |
| 41 to 45 | 73 | 3.29% |
| 46 to 50 | 45 | 2.03% |
| 51 and over | 46 | 2.08% |
| **Total** | **2,216** | **100.01%** |

For the entire (male and female) university student population in general per the registrar, the demographic makeup (2021) was Native American (0.11%), Asian (8.8%), Black/African American (17.6%), Hispanic (32.2%), Hawaiian or Pacific Islander (0.1%), Caucasian (29.7%), Mixed (3%), International (3.79%), and Unknown (4.7%).^19^ The racial/ethnic distribution of the respondents (just the female respondents) to this survey is presented in Table 3 and Figure 2.

**Figure 2.**
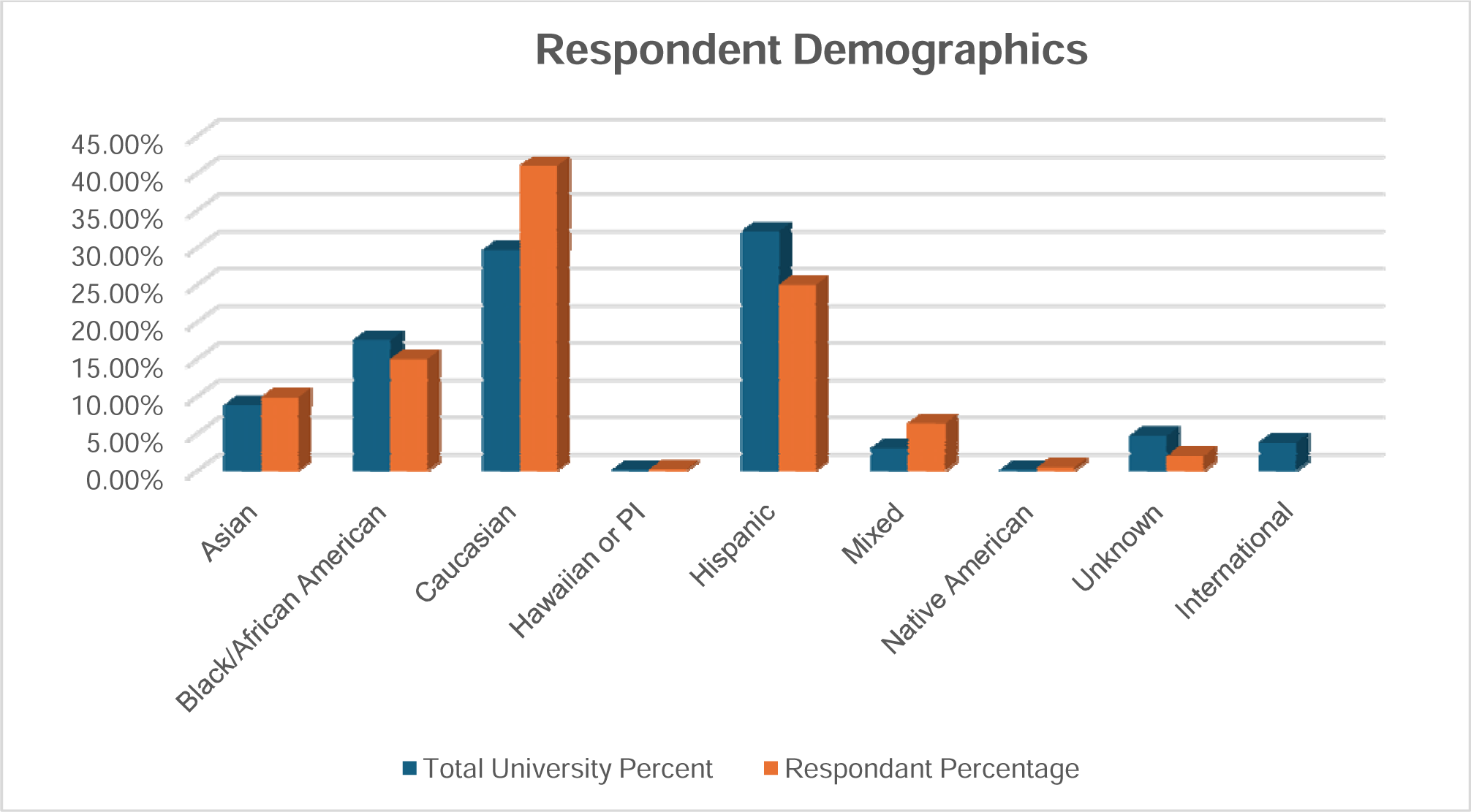
Respondent demographic percentages. There is an overrepresentation of Caucasian responses compared to the general student body (41% study vs 29.7% general) and an underrepresentation of Hispanic responses (25% study vs 32.2% general). There was no choice for international respondents in this study; 13 respondents did not answer this query out of 2,216.

**Table 3.** Racial/Ethnic Distribution. Respondent demographic percentages. There is an overrepresentation of Caucasian responses compared to the general student body (41% study vs 29.7% general) and an underrepresentation of Hispanic responses (25% study vs 32.2% general). There was no choice for international respondents in this study; 13 respondents did not answer this query out of 2,216.

| **Racial/ethnic group** | **Total University Percentage** | **Respondent Percentage** |
| --- | --- | --- |
| Asian | 8.80% | 9.90% |
| Black/African American | 17.60% | 15% |
| Caucasian | 29.70% | 41% |
| Hawaiian or PI | 0.10% | 0.20% |
| Hispanic | 32.20% | 25% |
| Mixed | 3% | 6.40% |
| Native American | 0.11% | 0.50% |
| Unknown | 4.70% | 2.00% |
| International | 3.79% |  |

### Menstrual Characteristics

The subjects reported normal cycling in 70.98% (1,573) of cases, irregular cycling in 21.30% (472), no cycling due to medications for endometriosis or oral contraceptives in 9.48% (210), no cycling due to previous gynecologic surgery (hysterectomy or ablation) in 1.04% (23), and no cycling due to menopause in 1.49% (33) of subjects. The top three symptoms reported by the study population were bloating/swelling/constipation (77.66%; 1,721), mood swings/moodiness (74.1%; 1,642), and pelvic or back pain (71.57%; 1,586). Of note is that 18.23% (404) reported bleeding that restricted the subject’s ability to function. The entire display is featured in Table 4.

**Table 4.**
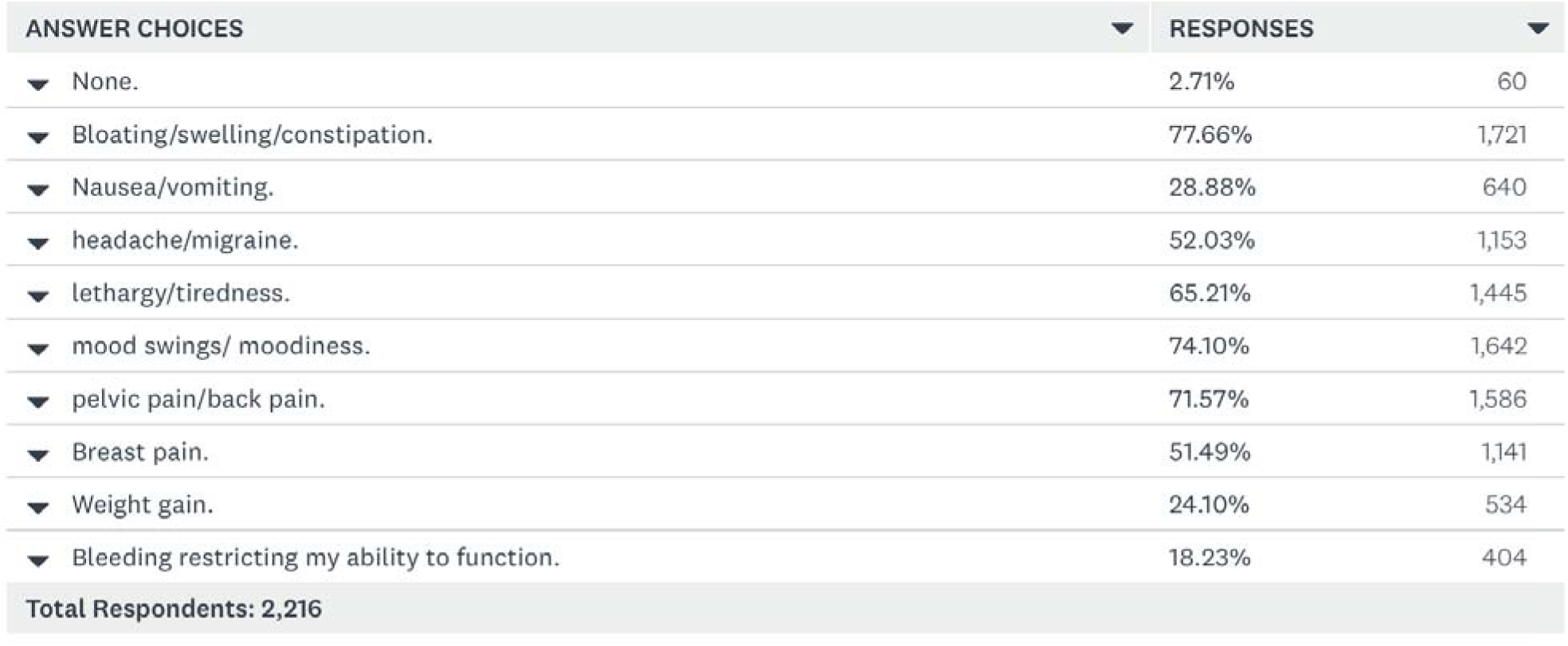
Menstrual Symptoms of the Respondents. The array of symptoms reported by respondents (more than one choice per person). Only 2.71% had no symptoms.

**Table 4.** Treatment Regimens for Menstrual Cycle Symptoms. Reported treatments used for menstrual symptoms in the cohort. 12.82% didn’t need anything, and only 1.99% used menstrual devices such as TENs or Jovi. (more than one choice was permitted). 79.29% used over-the-counter medications. The total number of respondents was 2,216; the percentages were established from the responses listed.

| ANSWER CHOICES ▼ | RESPONSES ▼ |  |
| --- | --- | --- |
| ▼ Nothing, I don't need treatment. | 12.82% | 284 |
| ▼ Prescription pain relievers | 8.35% | 185 |
| ▼ Over the counter pain relievers (Motrin, Advil, Aleve, Midol) | 79.29% | 1,757 |
| ▼ Over the counter menstrual pain devices (eg Livia, Jovi, TENS) | 1.99% | 44 |
| ▼ Oral Contraceptives/hormonal management | 25.09% | 556 |
| ▼ Cannabis/Marijuana/THC/CBD | 14.76% | 327 |
| ▼ alternative medicine/acupuncture/herbal medicine | 5.64% | 125 |
| ▼ Exercise/meditation | 29.60% | 656 |
| ▼ Alcohol | 2.21% | 49 |
| ▼ sexual relations | 9.12% | 202 |
| ▼ antidepressants | 3.16% | 70 |
| ▼ Heating Pad | 54.47% | 1,207 |
| Total Respondents: 2,216 |  |  |

When asked how severely their menses affect them, approximately 1 in 6 women (15.75%; 349) reported minimal to no issues. In contrast, 56.32% (1,248) had minimal to moderate issues that kept them from performing some of their daily routines, but they felt they could cope well. Almost 27% of the women (26.94%, 597) felt that their menses caused moderate to severe symptoms, keeping them from doing many things and interfering with their lifestyles. Severe symptoms that required the subjects to stay in bed and greatly restricted activities to cope were reported by 10.97% (243), and 2.26% (50) had unbearable symptoms and could not function during menses. Interestingly, 7.08% (157) were told that they had endometriosis by their doctor, by having surgery, or by a family member (not a physician). Fully 93.55% (2,073) had not been diagnosed with this disease across the entire cohort.

### Symptom Treatments and Efficacy

Numerous treatments have been used over the years to alleviate menstrual issues with varying success rates. In this survey, almost 80% used over-the-counter medications, and 12.82% did not need anything for treatment. The full responses are shown in Table 4 below.

One of the more important questions revolved around the perceived effectiveness of the various treatment categories respondents reported using in Table 4

1. Prescription pain medications: of the 185 respondents who use them, 32.4% found them very effective, 37.8% moderately effective, and 29.8% only mildly effective or ineffective.
2. Over-the-counter pain relievers – among the 1,757 respondents who use them, 27.3% found them very effective, 44.7% moderately effective, and 28% only mildly effective or ineffective.
3. Over-the-counter menstrual pain devices: of the 44 respondents who use them, 14% found them very effective, 33.2% moderately effective, and 52.7% only mildly effective or ineffective.
4. Oral contraceptives/hormonal management – of the 556 respondents who use this, 36.2% find it very effective, 33.6% find it moderately effective, and 30.2% find it only mildly effective or not effective at all.
5. Cannabis/Marijuana/THC/CBD – of the 327 respondents who use this, 51.7% find it very effective, 30.3% find it moderately effective, and 18% find it only mildly effective or not effective at all.
6. Alternative medicine/acupuncture/herbal medicine – of the 125 respondents who report using this, 20.3% find it very effective, 35.7% find it moderately effective, and 44% find it only mildly effective or ineffective.
7. Exercise/meditation: used by 656 respondents; of these, 17.5% find it very effective, 36.7% moderately effective, and 45.8% only mildly effective or not effective.
8. Alcohol – this is reported by 49 as a treatment, and of this group, 8.1% found it very effective, 11.5% found it moderately effective, and 80.3% found it only mildly effective or not effective at all.
9. Sexual relations – of the 202 respondents who engage in this as a treatment adjunct, 12.5% find it very effective, 32.8% find it moderately effective, and 54.7% find it only mildly effective or not effective at all.
10. Antidepressants are used as a treatment by 70 respondents, and of these, 5.9% found them very effective, 20.1% found them moderately effective, and 74% found them only mildly effective or not effective at all.
11. Heating pad – 1,207 respondents use this as a treatment for menstrual cycle symptoms. Of that, 24.5% found it very effective, 41.7% found it moderately effective, and 33.8% found it minimally effective or ineffective.

### Cannabis/marijuana

Of the 14.76% (327) of respondents who reported the use of cannabinoids for menstrual cycle symptoms, 89.91% (294) use smoking or vaping as a delivery system, 30.58% (100) use edibles/gummies, and 9.5% (31) use topicals, creams, or oils. Only one patient used a nasal or oral spray system, and one used a suppository system. In the next survey question related to cannabis/marihuana use, there were 412 respondents rather than 327. The “n” of 412 responses reflects that 18.6% of respondents used cannabis for menstrual symptoms, with 49.51% using conventional THC (delta 9 THC) dominant strains, 15.53% using a CBD (cannabidiol) dominant strain, 26.46% using an equal mix of THC and CBD strain (50/50 mix), and 8.5% using the delta 8 TCH products, with the rest of respondents stating that they did not use cannabinoids. The direct question of what was used seems to have engendered more responses. Recently, there have been reports of statistically significant pain reduction using cannabidiol (CBD) infused tampons to control dysmenorrhea, though the literature is sparse on this option.^20^

The respondents reported on the perceived effectiveness of the cannabis delivery systems in Figure 4.

**Figure 3.**
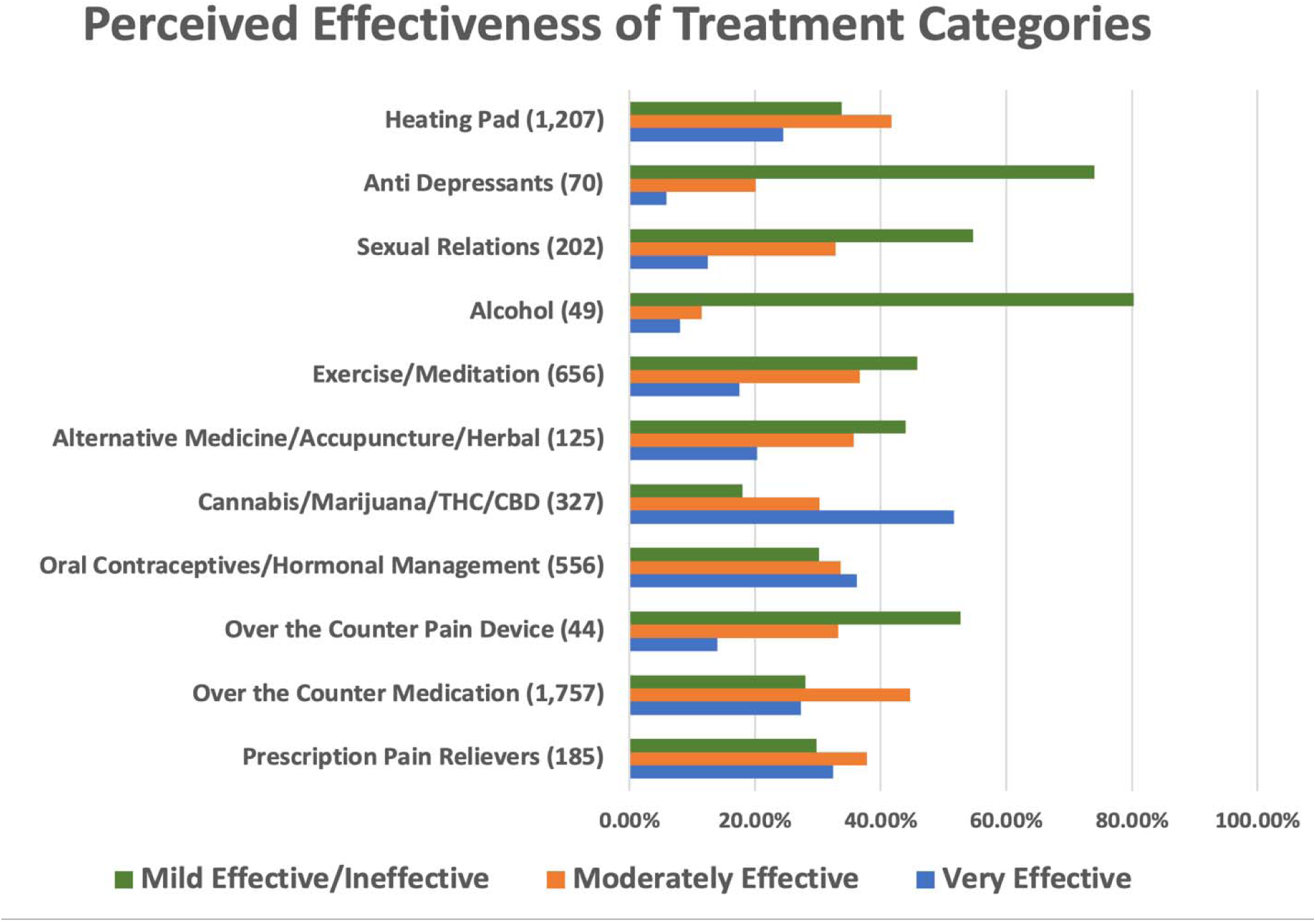

**Figure 4:**
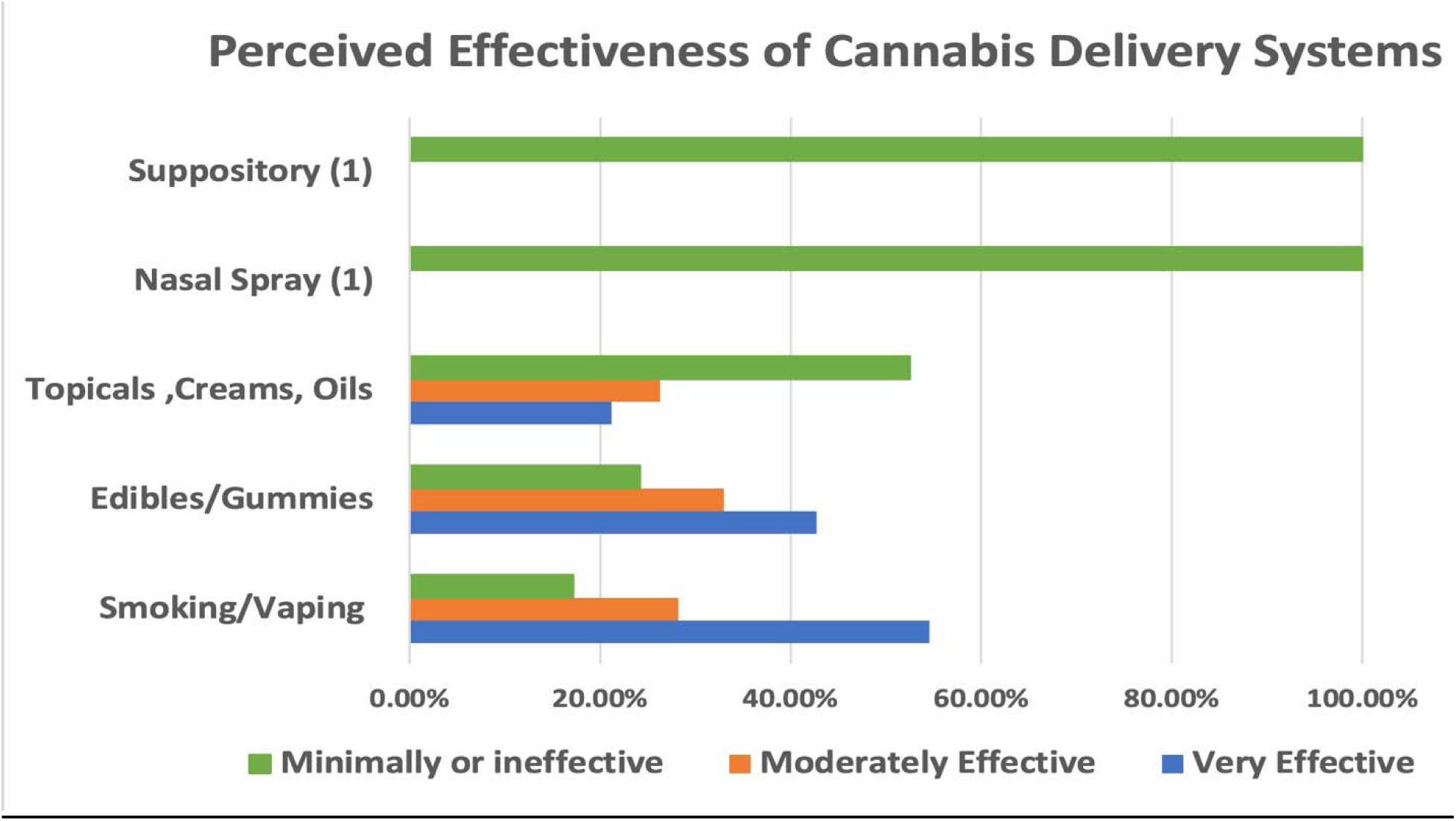

### Racial/ethnic differences

Some racial differences in responses regarding treatments were noted. The respondent’s age range did not appear to affect their responses.

1. **Black/African American** – A greater number of respondents did not need treatment for menstrual symptoms (16.92% vs. a mean of 12.82% for all other races/ethnicities), and a greater number required prescription pain relievers (15.11% vs. a mean of 8.35% for all other races/ethnicities). Further, compared to all other races and ethnicities, Black/African Americans reported less usage of oral contraceptives (13.9% vs. a mean of 25.9%), cannabis (8.76% vs. a mean of 14.76%), and exercise/meditation (20.54% vs. a mean of 29.6%).
2. **Caucasian** – There was a greater number of respondents using oral contraceptives (33.0% vs. a mean of 25.9% for all other races), and no significant differences for other treatments used compared to other racial/ethnic groups.
3. **Hispanic** – There was a lower number using antidepressants (1.44% vs. a mean of 3.16% for all other races), with similar usage of other treatments compared to all other groups.
4. **Asian** – Similarly, fewer respondents used antidepressants (1.83% vs. a mean of 3.16% (all other races)), with different treatments showing similar usage patterns compared to other races/ethnicities.
5. **Mixed** – those with mixed racial ancestry reported more use of cannabis (23.57%) compared to a mean of 14.76% for all other groups, and more use of exercise and meditation (38.57%) vs. a mean of 29.6% for all other groups.

## Discussion

For the female population, the burdens of menstruation, including symptoms of pelvic pain and psychological distress, are widespread, significantly impacting the quality of life, finances, education, and ability to work and function. Though previous studies have used social media channels, there is an inherent bias in recruiting through self-help channels. In this study, we included all individuals who identified as female from the entire university population to address this bias. Further, by including those with endometriosis, we tried to evaluate the disease burden and the various coping mechanisms used by this young female sample to alleviate their symptomatology. The survey response was 15.8%, similar to the standard response rate for other NSU university surveys. The study was weighted toward younger subjects, as might be expected in a university study, with 63.95% of participants aged 18 to 25. Demographically, Caucasians were overrepresented as respondents compared to the general student body (41% vs. 29.7%), and Hispanics were underrepresented (25% vs. 32.2%).

It was noted that 18.23% of the university’s female population reports pain and bleeding that restricts the student’s ability to function, and 13.2% were so severely affected by symptoms that bed rest was required, impacting academics and advancement, which has significant policy implications. Recently released guidelines on cannabis use throughout women’s lifespans from the Society of Obstetricians and Gynecologists Canada (SOGC) focused on several essential statements on cannabis use in the female population. ^21,22^ Canada legalized cannabis for recreational use countrywide on October 17, 2018 (Cannabis Act), and they have now had experience with large swaths of the population using it over time.^23^ This prompted the SOGC to issue its recommendations, shown below, with its confidence in the scientific evidence supporting each statement.

1. Cannabis use is increasing, and women are commonly using cannabis for recreational and medical reasons (high confidence). In the US, for the first time in 2023, 19- to 30-year-old female respondents reported a higher prevalence of past-year cannabis use than male respondents in the same age group, reflecting a reversal of the gap between sexes. Conversely, male respondents 35 to 50 years old maintained a higher prevalence of past-year cannabis use than female respondents of the same age group, consistent with what’s been observed for the past decade. ^24^
2. The use of cannabis, especially products containing tetrahydrocannabinol, can induce or worsen psychosis and depression (high confidence).
3. Study results concerning the association between cannabis and anxiety are conflicting, and this association requires further study. (High confidence).
4. There is limited evidence that frequent cannabis use may affect female fertility (moderate confidence), but frequent use can diminish male fertility (moderate confidence). A recent study by Duval et al. (2025) revealed significant effects on oocyte maturation, transcriptomic profiles, meiotic spindle organization, and oocyte ploidy. Collectively, this data presents compelling evidence that cannabis consumption may negatively impact female fertility.^25^
5. No evidence exists that cannabis products improve chronic pelvic pain (low confidence).
6. The effects of prenatal cannabis exposure on long-term outcomes through childhood, adolescence, and adulthood have not been conclusively defined. Still, recent data suggest that there are persistent neurocognitive effects into adulthood (moderate).

Our study seems to support findings of increased use of cannabis for self-treatment of menstrual symptoms, at least in the university female population we sampled. Moreover, Cannabis/Marijuana/THC/CBD had the highest perceived level of effectiveness against these symptoms among female users compared to other traditional treatments such as over-the-counter medications and oral contraceptives. This justifies further research into possible mechanisms that may explain this perceived benefit in a recurrent condition that females face regularly throughout their reproductive lives. Implications of cannabis use for women’s health, fertility, and its impact on offspring’s health must also be investigated.

## Conclusion

Menstruation and its associated symptoms in this college female population represent a significant challenge. Even though we did not evaluate the impact of menstrual burden on academic performance, the fact that 40.2% of the female college population had a significant impairment in their ability to function, with 13.23% being incapacitated entirely, makes menstruation a significant barrier compromising university life. Various adjuvants include the array of self-treatment modalities used by this population for symptom relief, including reliance primarily on over-the-counter medications for a substantial percentage of respondents with varying perceived efficacies. Additionally, cannabis is used by up to 18.6% as an adjunct with 80% perceived efficacy, deserving further studies. In addition to this study, the previous data cited on the effects of menstrual burden on academics, advancement, and quality of life should inform a policy review of how menstrual burden is addressed in university settings, particularly regarding efforts toward gender equality. The prevalence of cannabis use by nearly one in 5 students for menstrual burden is a reality that may elicit a policy review in many institutions, both at the university and secondary school levels. An awareness of the extent of the burden may help adjust testing schedules and teaching formats to accommodate this large portion of our population.

### Advantages

The entire university body that identified as female was invited to participate, regardless of social media platforms and the biases they entail.

### Limitations

The 15.8% response rate is disappointing, though that rate (about 16%) is commonly encountered in this university’s surveys, and the 2,216 responses are certainly one of the larger ones published on this subject. Response bias is a possibility, as in all studies, and the cited placebo effects may compromise questions on cannabis and other treatment modalities.

## Data Availability

All data produced in the present work are contained in the manuscript

## Notes

### Competing Interest Statement

The authors have declared no competing interest.

### Funding Statement

This study did not receive any funding

### Author Declarations

The study was approved by the Nova Southeastern University Institutional Review Board (Study on Women's Menstrual Symptoms and Treatment NSU IRB Protocol Number 2021,544).

